# Use of Packed Red Blood Cell Mechanical Fragility to Indicate Transfusion Outcomes

**DOI:** 10.1101/2022.05.28.22275705

**Authors:** M. Tarasev, S. Chakraborty, K. Alfano, M. Muchnik, X. Gao, R. Davenport

## Abstract

The hypothesis for this study was that RBC mechanical fragility (MF) could be an aggregate *in vitro* property predictive of transfused RBC performance *in vivo*. Various MF values were obtained via MF profiling, based on several variations of testing parameters, using both a “legacy” approach (with a commercial, cam-based vertical bead mill and a spectrophotometer) and a more proprietary approach (with a custom-developed, electromagnetic horizontal bead mill combined with proprietary optics and analysis). A total of 52 transfusion events in 32 different patients recruited from the University of Michigan were included in this study. Results were assessed using mixed effects and linear regression models. RBC MF was shown to predict about 15% of transfusion-associated changes in patient hemoglobin concentration, but not of secondary hemolysis-associated metrics (serum hemoglobin, HAP, and LDH). This result was affected by several factors that were not fully accounted for, including variability in post-transfusion blood collection time and variability in each blood unit volumes. Inclusion of the number of units transfused showed the potential to improve predictive capability, thus highlighting the potential importance of underlying patient condition necessitating the second unit transfusion. Certain ways of applying the bead-induced mechanical stress showed MF results more suitable for predicting transfusion outcomes than others indicating potential significance of flow stress type for assessing storage-induced RBC membrane damage. That highlights an opportunity for improvement of the potential for use of MF metrics, through identification of optimal stress application parameters (possibly by further varying parameters used here, as well as others) for assessing contribution of storage-lesion-associated RBC damage on transfused RBC performance.

## 1. Introduction

Unlike chemical-based drugs, biological products such as packed Red Blood Cells (pRBC) cannot be easily standardized, as the nature and properties of cells to be transfused varies at the time of blood collection, with differences further amplified through blood processing and pRBC storage. The NIH 2016 meeting on Scientific Priorities in Pediatric Transfusion Medicine set up priorities for development transfusion therapy practice, including “developing new methods for better blood management and transfusion decision,” and “determination of packed Red Blood Cell (pRBC) characteristics that impact RBC survival, function, and clearance in chronic transfusion patients” [1]. This dovetails with the one of the highest scientific priorities identified in the proceeding of NIH/NHLBI State of the Science in Transfusion Medicine Symposium, “Can the potency and/or safety of transfusable RBCs be improved?” or simply, “What is in the red blood cell bag?” [2, 3].

FDA regulations require < 1% pRBC hemolysis at the end of storage, and a mean 75% cell recovery with a one-sided lower confidence limit of 70% at 24 hours post-transfusion, thus allowing for ∼10% of all pRBC units to have below the 75% recovery. New products are validated by reinfusion of autologous ^51^Cr-labeled RBC at the limit of storage to healthy volunteers, with the results then extrapolated to the over 13 million pRBC units annually manufactured and transfused in the Unites States [4].

RBC viability varies significantly at the end of storage, with recoveries ranging from below 60% to over 95% (e.g. [5]). Dumont and AuBuchon [6] reported that while the majority of units had recoveries of 75-80%, consistent with the FDA benchmark, some units had recoveries close to 100% while others had 55-65% recovery rates – with the minimum observed recovery value being 36%. As was pointed out, when extrapolated to ∼13 million units annually transfused, the number of such units with unacceptably low recovery could be alarmingly large. Moreover, 75% recovery implies that a quarter of transfused cells are not contributing to oxygen delivery, and in cases of blood exchange, for example, adds to iron overload. While manufacturing plays a role [7-14], a growing consensus is that the variability in post-storage pRBC viability, in terms of both *in-vivo* survival and oxygen delivery, is primarily due to donor variation rather than variance in storage conditions [15, 16].

Physiological removal of transfused RBCs from circulation is a non-linear process [17]. A fraction of cells (0-15%) – possibly representing senescent, highly damaged, and more rigid RBCs – is removed shortly (< 1h) post-transfusion [18, Barshtein, 2017 #47759, 19]. Additional rigid and fragile RBCs are lost over the next several hours (e.g., reaching ∼22% total at 4 hours [20]), likely due to shear forces in the bloodstream, with the “fast” clearance essentially complete after 24 hours [21, 22]. Remaining cells (∼75% of those transfused) remain in circulation subject to 0.8-2% per day cell loss. Reports of longer-term removal/survival include about 20% of transfused cells surviving after 32 days [20], and 50% at 30 days [23, 24]. Notably, there could be significant differences between individual recipients as well (e.g., [25, 26].

Underlying causes of the variability in progressive biochemical and morphological changes which occur during RBC storage, collectively known as “storage lesion,” have been extensively studied and described [27, 28]. The changes involve a wide range of highly integrated cellular parameters and functions [29] that impact cell viability, oxygen delivery, and hemodynamics – all of which affect transfusion outcomes (see [29] for likely clinical sequelae). Some of these changes are reversible in vivo, with cell recovery times varying from several hours to several days [30, 31]). Such recovery involves delays and biological costs, and its significance would often be different for different recipients (e.g., those with acute need vs. those routinely transfused for chronic conditions).

While the use of pRBC time in storage as predictor of transfusion outcomes entails some controversy (see [32] for a review), there has been interest in identifying other indicators of potential Red Cell post-transfusion performance [33]. Comprehensive answers should account for both donor and recipient variability, with the question “is older blood bad?” not being “oversimplified to the point of being neither testable nor applicable” [34].

Donor variables contribute to stored pRBC unit variabilities including ATP levels [35-37], pRBC metabolic age [38, 39], “in-bag” hemolysis [8, 40], cell morphology [41], and cell lifespan (which has a mean value of 115 days, but exhibits individual variability in the range from 70 to 140 days) [16, 39]. Factors like ethnicity, age, gender, frequency of donation, lifestyle (e.g., diet, physical activity, etc.), pre-clinical medical conditions and genetic pathologies all likely to contribute [16, 42, 43]. Potentially hereditary genetic polymorphisms were shown to contribute to variability of RBC survival, both in storage and post-transfusion [44-46]. The question of the impact of donor variables was listed as a priority by the 2015 State of the Science in Transfusion Medicine symposium [3].

Although it remains to be shown which *in vitro* cell parameter(s) can be reliably correlated with transfusion efficacy and outcomes, mechanical properties of RBC membranes have been shown to be affected by *in vitro* storage [47-50], with changes in membrane functions relating to changes in RBC metabolic state [51, 52]. Mechanical fragility (MF) [50, 53] and related flow properties [54, 55] have been proposed as potential candidates to supplement storage time as an aggregate metric of pRBC storage lesion. MF had been much less explored, due in large part to the lack of a convenient and standardizable MF testing system [56]. Overall, differences in RBC functional properties due to things including manufacture of packed RBC units (pRBC), their storage, and performance when re-introduced into circulation, collectively determine pRBC recovery rates *in-vivo* [57]. That necessitates the development of testable functional markers predictive of transfusion recovery.

This study aimed to validate the use of various potential RBC MF metrics as such a marker, using simple transfusion performance evaluation based on post-transfusion hematocrit/total hemoglobin. That method was used previously by Pieracci et. al. [58], although in that application the accuracy of the results could potentially be improved by additional corrections for transfused unit hemoglobin content and for patient pre-transfusion blood volume. The advantages of the approach combine its ease of implementation and suitability to use with large subject numbers with the fact that patient hemoglobin remains one of the dominant indicators/triggers for RBC transfusion in clinical practice. Additionally, unlike most other methods, use of Hb/HCT as an outcome metric requires no modification of currently approved RBC products and has minimal impact on Standard of Care (SOC) transfusion procedures.

## Materials and Methods

The study had been conducted according to a protocol approved by the University of Michigan Institutional Review Board, which gave ethical approval for this work.

### Enrollment

A total of 41 different patients were identified and consented for participation in the study, with 21 being female and 20 being male. Age of participating subjects was 61 ± 16 years (mean ± SD), with the lowest age of 22 years and highest age of 83 years. For 3 patients, no sample was collected, due to changes in treatment. For 7 patients, samples of pRBC received post-transfusion could not be tested for RBC Mechanical Fragility, due either to sample contamination (e.g., with physiological solution), low collected volume, or excessive delay in transportation.

### Transfusion events

A total of 52 independent transfusion events were analyzed in the study, among 32 patients participating (14 females and 18 males) and receiving blood transfusion, and with a pRBC sample being analyzed through RBC MF assays. 10 patients received several transfusions over an extended period of time – with 5 patients participating twice (two independent transfusion events), 3 participating three times (three independent transfusion events), and 2 participating four times (four independent transfusion events). Out of the total of 52 transfusion events, 32 were single unit transfusions, and 14 involved two pRBC units. When two pRBC units were transfused, mechanical fragility was measured on a sample that represented a mix of pRBC from both units. Transfused pRBC was collected in CPD, CP2D, or CPDA1 and stored respectively in AS1, AS3, or CPDA1 storage solution. These units differed in pRBC volume, with Table 1 showing an average unit volume and standard deviation based on volume measurements (by weight, corrected for the weight of the empty bag and segments) of 30 units for each of the storage conditions. Differences between average volumes of three pRBC unit types (with three different storage solutions) were found to be statistically significant (with α <0.05). Measurement of volumes of actual units transfused was not possible due to operational constrains and the need to not disrupt standard operating procedures, thus an adjustment to the calculation was made based on average unit volumes. Imprecision of such an adjustment represents one of the limitations of this study.

**Table1.**
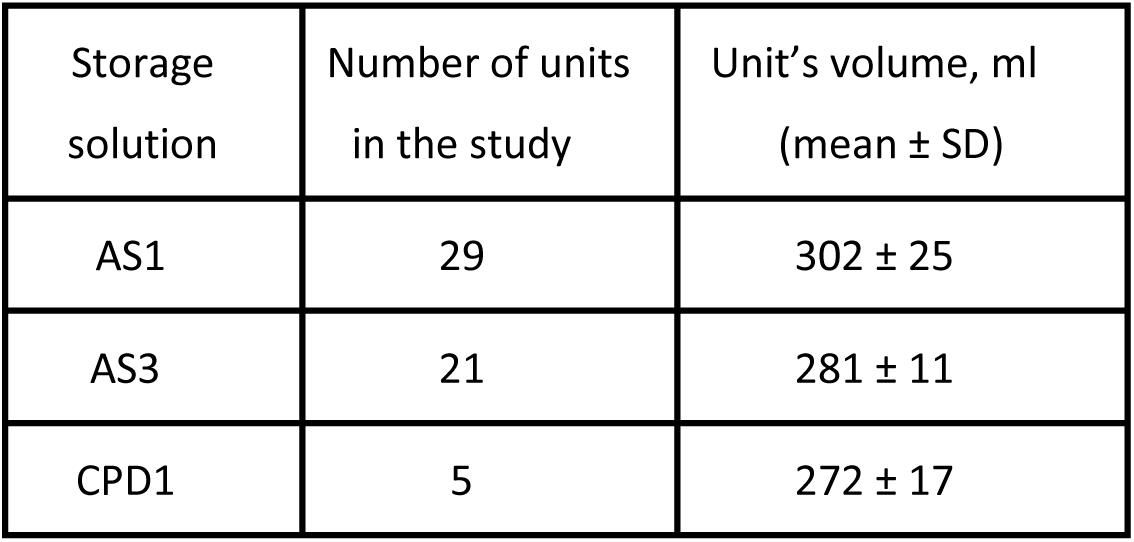
Volume of pRBC units with different storage solutions in the University of Michigan Blood Bank.

| Storage solution | Number of units in the study | Unit's volume, ml (mean ± SD) |
| --- | --- | --- |
| AS1 | 29 | 302 ± 25 |
| AS3 | 21 | 281 ± 11 |
| CPD1 | 5 | 272 ± 17 |

### Sample collection

Each sample for the fragility testing was obtained from the residual pRBC in the “empty” bag after transfusion. At the end of a transfusion, such an “empty” bag typically contains ∼2-3 ml of pRBC, volume sufficient for a range of Mechanical Fragility tests involving different conditions and different methods of stress application (described below). Samples for blood analysis before and after the transfusion event were collected when ordered by the attending physician and analyzed for hemoglobin (Hb) concentration (by CBC) and red cell hemolysis markers including lactate dehydrogenase (LDH), bilirubin (BUN), haptoglobin (Hp), and serum free hemoglobin (SFH). Blood collection in the study protocol was aimed at obtaining a post-transfusion blood sample as close as possible to 24 hours after the transfusion. While effort was made to follow this timing, some samples were collected with unrecorded variation in collection time, which contributes to random error in correlative analysis involving post-transfusion blood properties. Hemoglobin concentrations were verified using a Hemoglobin 201 system from HemoCue (Angelholm, Sweden).

### Mechanical stress application and induced-hemolysis profiling

Mechanical shear stress was applied through two different bead-milling methods, one using a vertical cam-based commercial system and the other based on a proprietary electromagnetic horizontal bead mill approach. These two stress application methods (described further below) involve different bead and sample tube geometries, necessitating the use of significantly higher oscillating frequencies in the cam-based system to achieve cumulative hemolysis comparable to that in the electromagnetic setup. Speed of the flow through the annulus of an oscillating cylindrical bead in the tube which is dependent on the speed of the bead and the size of the gap between the bead surface and the wall of the tube, has been shown to be determinatives of generated flow shear stress and resultant hemolysis [59]. Presence of the air with the sample in the cam-based system, as compared to the proprietary electromagnetic approach where no such is present, can potentially add additional stress to cells at the liquid-gas interface. Overall, it was anticipated that differences between the systems as well as more specific assay parameters may impact both the magnitudes and the types of mechanical stresses being generated, potentially providing different assessments of RBC lysis propensities.

#### “Legacy” cam-based system

This approach utilized a commercial cam-based vertical bead mill (Qiagen GmbH, Redwood City, USA) for stress application, combined with a spectrophotometric analysis of supernatants as described previously [54, 55], except with custom-made stainless steel cylindrical beads. Depending on their length and diameter, cylindrical beads were previously shown to generate categorically different types of mechanical stress significantly dependent on erythrocyte environment [59]. Bead dimensions (7 mm in diameter and 18 mm in length) used in this work had been selected so as to provide a substantial difference in induced hemolysis between samples supplemented and non-supplemented with albumin (SA; Bovine Serum Albumin; RPI Corp, Mt. Prospect, IL). Oscillation at 20 Hz frequency was selected to provide nearly full hemolysis in samples not supplemented with SA within 20 minutes of stress duration. In SA supplemented samples, where at 20 Hz the hemolysis is very low, an increased (30 Hz) oscillation frequency was used instead. As hemolysis dependence on frequency of oscillation can be non-linear, in SA supplemented samples an additional measurement at 5 minutes of stress application at 20 Hz was conducted as a control and for direct comparison of the results. Prior to analysis, pRBC samples were diluted to 1.2 g/dL with Additive Solution 3 (AS3), pH 5.8, containing as necessary 4 g/dL bovine serum albumin (BSA; RPI Corp., Mt Prospect, IL).

#### Proprietary electromagnetic system

The approach utilized a proprietary electromagnetic horizontal bead mill, combined with a proprietary spectral analysis method that allowed noninvasive determination of hemolysis in the sample within the cuvette/tubing as described previously [60]. A ferromagnetic bead with a biocompatible black epoxy coating was oscillating within a sample containing flexible Tygon™ tube. “Short” (MF-S) and “Long” (MF-L) tube/bead configurations were used to provide qualitatively different mechanical stresses arising from differences in RBC suspension flow around the cylindrical beads. “Short” configuration used a tube of 4.85 mm x 34 mm with a 3.7 mm x 7 mm bead oscillating at 10 Hz, and the “Long” configuration used a tube of 4.85mm x 44 mm with a 3.7 mm x 18 mm bead oscillating at 5 Hz (dimensions are diameter x length). Only MF-L was measured on samples both supplemented and non-supplemented with BSA, as prior work showed MF-S to be only minimally affected by BSA supplementation. Prior to analysis, pRBC samples were diluted to 0.5 g/dl hemoglobin concentration with AS-3, pH 5.8, containing serum albumin when indicated.

Control experiments demonstrated that test results did not depend on sample hematocrit within a 0.4-g/dL range, with readings being within the dynamic range of the detection system.

### Hemolysis Assessment

Hemolysis (Hem), both present in untreated sample and induced by bead milling, was expressed as a fraction of cell-free hemoglobin (Hb^F^) relative to total hemoglobin concentration (Hb^T^) according to Formula 1 which included the correction for sample hematocrit as detailed by Sowemino-Coker [61].

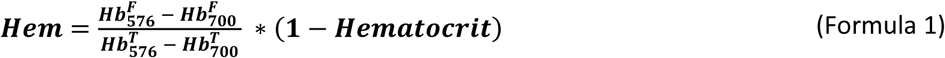

Total hemoglobin concentration for each diluted RBC sample was determined by subjecting a small (350-400 µL) aliquot to ultrasound for 40 seconds (0.1 second pulses, with 0.2 intervals between pulses, on ice) provided by a Branson Digital Sonifier 450 (Danbury, CT), at 15% intensity (from the manufacturer-specified 400 Watt). In control experiments, such treatment was shown to fully lyse RBC without inducing hemoglobin oxidation. Standard spectroscopic measurements were performed with a NanoDrop N1000 spectrophotometer (Thermo Scientific, Waltham, MA). Cell-free hemoglobin fraction was determined non-invasively within the tube, after each respective applied stress duration, using a proprietary spectral analysis approach as detailed previously [60, 62].

**RBC fragility profiles** are defined by a sample’s incremental hemolysis levels resulting from applied mechanical stress of varying (and here, successively cumulative) durations. Unlike single-point measurements that use a single stress duration at a single stress intensity, as implemented for example by Raval et al. [50], MF profiles allow recording RBC propensity to hemolyze over the range of applied stress magnitudes – from that resulting in minimal hemolysis to that resulting in nearly total hemolysis of cells in the tested sample – thereby allowing multiple fragility-based indexes to be interpolated for separate analyses [53]. Here, profiles are described by hemolysis parameters (Hem-parameters) representing the amount of hemolysis achieved at a given stress intensity in a given environment as a result of small (3 min) and large (10 min) stress durations, which are identified by a subscript number for the Hem parameter. Fragility parameters for each stress duration were obtained from best fit second-order polynomial regression to the experimental data. For curves exhibiting significant deviations from a simple polynomial, raw data was subdivided into low and high hemolysis sub-sets and the fits were obtained independently for each subset of the data.

### Outcome metrics

#### Primary outcome metric

(VARHBAD) represented the difference between blood hemoglobin concentration achieved after the transfusion and the theoretical blood hemoglobin concentration calculated based on the subject’s pre-transfusion hemoglobin and hemoglobin amount delivered through pRBC transfusion. Thus, it describes the “deviation” of the clinical outcome from that which would have been anticipated, potentially reflecting a “fast” (occurring in under 24 hours post-transfusion) phase of pRBC *in-vivo* lysis. Subject pre-transfusion blood volume was estimated as described by Feldschuh et. al. [63]. Transfused volume was estimated based on the average pRBC unit volume of an appropriate type, as given in Table 1. The metric was calculated using Formula 2 here:

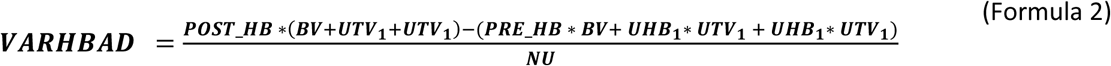

Where POST_HB and PRE_HB is subject hemoglobin measured before and after transfusion, BV is the subject’s estimated blood volume, UTV1 and UTV2 represent volumes, UHB1 and UHB1 represent hemoglobin concentration of the first and second transfused units respectively, and NU is the number of units transfused (either 1 or 2).

#### Secondary outcome metrics

included changes (post-transfusion value minus pre-transfusion value, normalized to the number of units transfused) in haptoglobin (D_HAP), lactase dioxygenase (D_LDH), total patient hemoglobin (D_HB), and serum hemoglobin (D_S_HB). Changes in haptoglobin, serum Hb, and LDH were expected to be partially correlated, with positive correlation anticipated between D_S_HB and D_LDH, and negative between D_S_HB and D_LDH with D_HAP.

Additionally collected were certain patient demographics (age, gender) and medical data (body mass index, blood type), and transfused pRBC unit information including blood type, storage solution, time in storage, and in-bag pre-existing hemolysis.

### Statistical Analysis

In this study, some patients were included more than once. Although the individuals themselves are assumed to be independent of each other, observations obtained from the same subject are correlated by sharing characteristics. Such data was described through linear mixed model. Typically, mixed effects models describe the population average regression model of outcome Y over a range of predictors (or fixed effects), plus subject-specific components (or random effects). Predictors considered for this model were varied to include demographics, pre-transfusion patient metrics, and transfused unit properties including their mechanical fragility (MF) metrics. The subject-specific components used in this model were random intercept and random slope, over one or more predictors. The analysis was performed using the statistics software R, version 4·0. (R Foundation for Statistical Computing, Vienna, Austria).

Linear regression with ANOVA was used, as appropriate, with datasets containing repeat measurements subdivided into multiple datasets of single measurement data, independently analyzed and compared for variance. Data is presented in terms of mean ± SD, with distribution as shown using a Box-and-Whisker’s plot, with the box showing upper and lower quartiles, the median, and the mean (X), and lower and upper whiskers positioned at 1.5 x IQR (interquartile range), with data points (including outliers) shown. Statistical significance was set at p<0.05.

## Results

### Transfusion-associated changes in outcome metrics

Total patient hemoglobin would be expected to increase after transfusion, with such increase proportional to the pRBC recovery rate after 24 hours following the transfusion event. In cases where all RBC survive right after the transfusion and there is no new RBC generation (a reasonable assumption for the 24-hour time frame), a VARHBAD value of zero would be anticipated. Similarly, zero changes would be anticipated for D_LHD, D_HAP and D_S_HB – reflecting the absence of transfusion-associated hemolysis. Post-transfusion pRBC degradation and hemolysis would be associated with negative values of VARHBAD, increased D_LDH and D_S_Hb, and decreased D_HAP. Observed distributions of the values of outcome metrics are presented in Figure 1 as Box-and-Whiskers plots.

**Figure 1.**
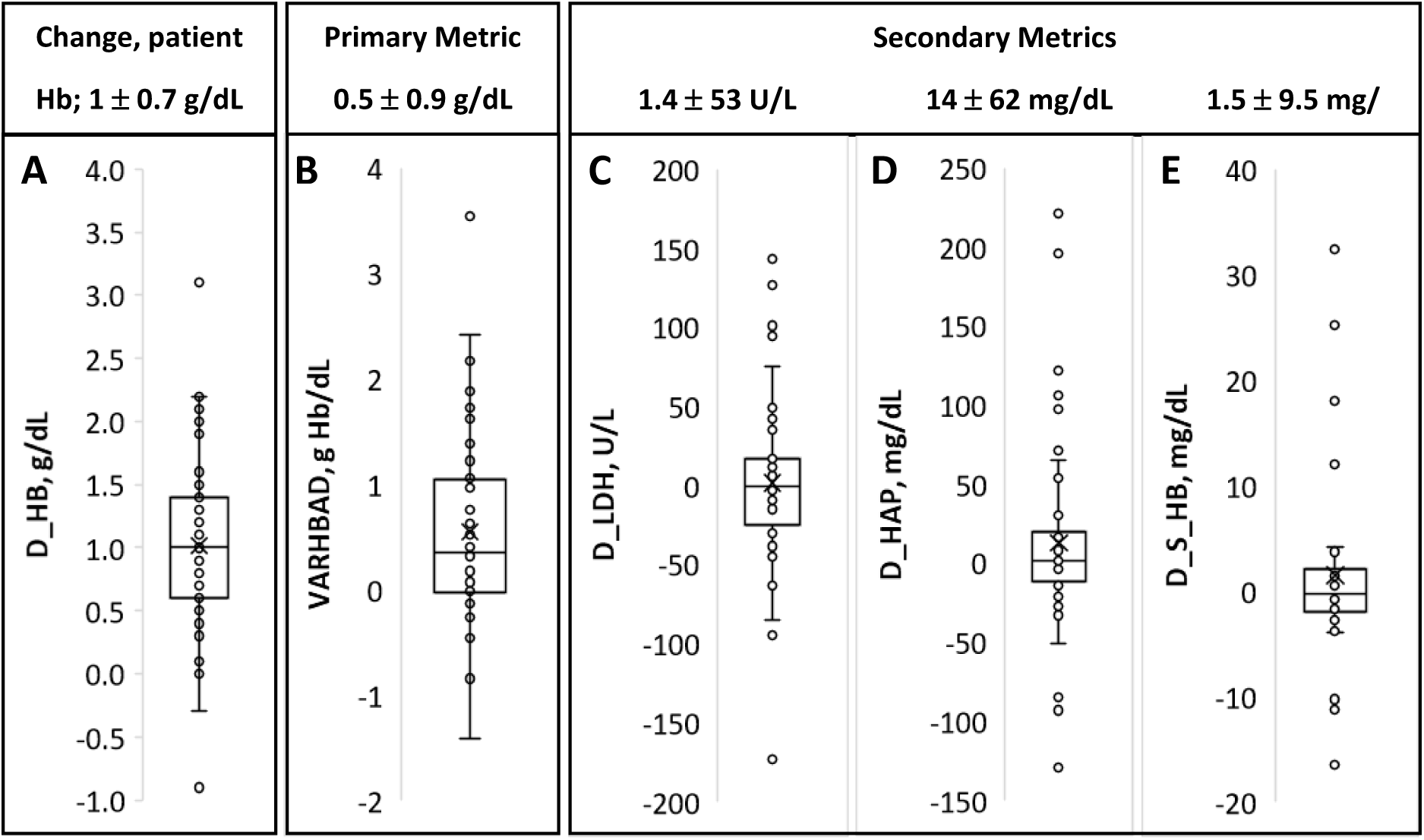
Distribution of outcome metrics. VARHBAD (A), D_LDH (B), D_HAP (C), D-S-Hb (D) and D_Hb (E). All changes are per pRBC unit transfused. The box represents Q1 to Q3 interquartile range with a horizontal line in the middle to denote the median. Mean of the data set is denoted as X. Boundaries of the whiskers correspond to the minimum and maximum values of the data set (excluding any outliers).

As measured, change in patient Hb varied significantly between transfusion events (Figure 1D), with an average observed increase of about 1 g/dL per pRBC unit transfused typically considered a good transfusion outcome (Figure 1A). However, despite transfusions’ seemingly achieved therapy goals, transfusion-related increase in patient Hb was consistently lower than that anticipated based on the pRBC volumes and hemoglobin content. The average VARHBAD value, across all transfusion events, was 0.54 ± 0.88 and was highly varied between the events with the range from 2.5 to negative 1.4 (excluding the outlier; Figure 1B).

While average (over all transfusion events) changes in the three metrics used to estimate *in-vivo* RBC hemolysis were relatively small, with median values at about zero, all metrics exhibited high variability between transfusion events (Figures 1C-1E). In this study, no significant correlations between these metrics were observed: changes in LDH concentration were not correlated with changes in serum Hb or HAP concentrations, and there was no correlation between the observed changes in serum Hb and HAP.

### Patient attributes

Patient’s age, gender, body mass index, or pre-transfusion blood volume were not predictive of either the primary transfusion outcome, VARHBAD, D-HB, or the secondary transfusion outcomes D_LHD, D_HAP, or D_S_HB.

### pRBC unit properties

A total of 65 units had been transfused in the study. The distribution of parameters associated with these units is given in Table 2.

**Table 2.** Properties of the transfused pRBC units

| Total units | Blood type |  |  |  | Rh factor, %Rh<sup>+</sup> | Storage solution |  |  | 2 units used | Irr.* | UHB, g/dL | AH, % |
| --- | --- | --- | --- | --- | --- | --- | --- | --- | --- | --- | --- | --- |
|  | A | B | O | AB |  | AS1 | AS3 | CPDA |  |  |  |  |
| 65 | 20 | 4 | 40 | 1 | 75% | 34 | 26 | 5 | 14 | 13 | 18.5 ± 3.3 | 2.8 ± 1.8 |

\* Irr. – number of irradiated units

Notably, units’ measured pre-existing hemolysis values (auto-hemolysis, AH), were significantly elevated as compared to industry standard of 1 percent maximum in the US. Those values, however, do not necessarily reflect pRBC auto-hemolysis at the time of transfusion but also additional lysis due to transfusion-associated activity from the procedure, as well as extra storage and handling associated with harvesting residual sample from the “empty” bags.

Mixed effect model analysis did not show a correlation between AH and primary or secondary outcomes metrics. No correlation was found with the unit hemoglobin concentration. The ABO blood group and irradiation status also were not predictors of either primary or secondary transfusion outcomes. Potential impact of the Rh factor could not be evaluated due to its skewed distribution (see Table 2).

Average storage time of the pRBC units used in this study was 19 ± 8 days, ranging from 3 to 39 days. For transfusion events where a single pRBC unit was used, storage time was not a significant predictor of primary or secondary outcome metrics. Including in the model transfusion events with 2 units transfused makes storage time a significant predictor of the primary outcome. However, there is no apparent physiological difference to suggest why storage time would become a predictor in 2-unit transfusions only. Moreover, in most 2-unit events (62%), storage time for the two units was within 3 days of each other, and exceeded 10 days in one case only. At the same time, the number of units transfused was also found to be a significant predictor (p<0.05) of primary outcome, about 25% predictive towards VARHBAD. Thus, it seems possible that dependence on the storage time observed when 2-unit transfusions were included in the analysis could reflect a confounding effect of the two units (as opposed to a single unit) being transfused, and thus of the underlying correlation to the number of units transfused. That, in turn, could be also confounded with the underlying patient’s condition that required the transfusion of a second unit. However, the reliability of this inference is limited due to the limited number of the relevant observations.

Similar limitation applies to the use of storage time as a predictive metric for secondary transfusion outcomes in a mixed effect model, even further limited due to an even lower number of observations for these metrics. Storage time was not a reliable predictor of hemolysis-associated transfusion outcomes D_LDH, D_HAP, and D_S_HB.

### Pre-transfusion blood chemistry tests

Several pre-transfusion patient blood chemistry parameters had been evaluated as potential predictive variables for primary and secondary transfusion outcomes. No correlation was observed between the pre-transfusion levels of patient Hb, serum Hb, haptoglobin, or LDH and the primary outcome metric VARHBAD.

Post-transfusion values of hemolysis-related biomarkers LDH, HAP and serum Hb showed statistically significant correlation with their pre-transfusion levels. Specifically, pre-transfusion values explained about 17%, 25%, and 35% of variance in post-transfusion values of serum Hb, haptoglobin, and LDH correspondingly. However, the correlation between pre- and post-transfusion values of total patient Hb lacked significance. Correlation of pre-transfusion values of these biomarkers with the magnitude of the change in the biomarker value (post-transfusion value minus pre-transfusion value), with and without normalization to the number of pRBC units transfused, was much weaker for LDH (R^2^ =0.15) and lacked significance for patient Hb, serum Hb, and HAP. This observation supports the association of changes in hemolysis-related parameters with the transfusion event, as opposed to the pre-transfusion patient condition. Lack of significance in correlation with post-transfusion values observed for patient total Hb likely reflects stronger dependence of hemoglobin levels on transfusion than that for evaluated hemolysis biomarkers.

### pRBC Mechanical Fragility

#### “Legacy” cam-based system

When the mechanical stress was applied using the noted cam-based bead milling approach, measured mechanical fragility (MF-CAM) was not found to be a predictor of either primary or secondary outcome metrics. Significance was lacking regardless of bead oscillation frequency, or the sample being supplemented or not supplemented with BSA.

#### Proprietary electromagnetic system

With the other overall approach, three different stress application regimes/configurations (Short, Long, and Long supplemented with albumin; see Materials and Methods) had been preselected in an attempt to better match applied stress to that most relevant for storage-induced pRBC damage and post-transfusion survival. No significant correlation was observed between MF and primary or secondary outcome metrics when samples were diluted with AS3 supplemented with BSA. However, mixed effect model with repeat measurements shows a statistically significant correlation between MF metrics and transfusion-induced changes in hemoglobin when measured in pRBC samples with no BSA supplementation. MF expressed as Area Under the Curve (AUC) of the best fit to the experimental data (percent hemolysis over the duration of stress application) showed about 15% predictive capability towards VARHBAD for the Long configuration when tested with no BSA supplementation. While still statistically significant (p < 0.05), a similar correlation with MF-S had a smaller explanatory significance towards VARHBAD (R^2^ of about 0.05). A multiparametric model – including, in addition to MF-L, also the number of pRBC units transfused (another independent predictor of VARHBAD), allowed prediction of about 30% of the VARHBAD variance (with model significance F < 0.01, and both predictors being independently significant).

MF measured under both configurations had no significance as a predictor of transfusion-associated changes in LDH and HAP; however, MF measured using L configuration with no albumin was found to be a strong predictor of changes in serum Hb (p<0.05; R^2^=0.42).

Mixed effect models also showed change in LDH to be negatively correlated with the primary outcome (p<0.01, R^2^=12%), with unexpectedly an even stronger negative correlation with post-transfusion LDH levels (p< 0.01; R^2^ = 19%). No similar correlation was observed for serum Hb or HAP.

## Discussion

The transfusion of one pRBC unit is typically expected to increase patient Hb by about 1 g/dL. Research studies, however, report smaller and highly variable changes in patient Hb as a result of blood transfusion [19, 58]. Notably, Berenstain et. al. [19] observed such smaller than anticipated hemoglobin increases after transfusion even in splenectomized patients, where the clearance of storage-modified and then transfused RBC is expected to be slower. The study presented here also reports significant variability between transfusion outcomes, as expressed by the difference between achieved and anticipated transfusion-caused increases in patient Hb. While the mean value of Hb increase was indeed close to 1 g/dL, individual values ranged from negative 0.25 to positive 2.5 g/dL per pRBC unit transfused (Figure 1A). Notably, no correlation was observed between this variability and patient weight, BMI, or gender.

RBC hemolysis results in the release of LDH into the medium along with hemoglobin and its degradation products, which are then scavenged by body clearance and detoxifying systems. Correlation of LDH, both as measured post-transfusion and the magnitude of transfusion-associated change in concentrations, with change in hemoglobin would support hemolysis as potential root cause of Hb being lower after the transfusion then anticipated. Lack of similar correlations with serum Hb and HAP could then be interpreted as the hemolysis level being low enough not to overwhelm the Hb scavenging systems.

It was reported previously that transfusion outcomes, defined as change in patient Hb, showed significant positive correlation with pRBC deformability. Specifically, transfusion-associated increase in Hb was best correlated with the percent of low-deformability cells in the pRBC population (p = 0.0006, R^2^=0.23) [19]. That work also reported that pRBC with a low level of rigid RBC yields a longer interval between two consecutive transfusions, suggesting that “better” RBC, likely with less accumulated membrane damage, would endure longer in the circulation. Clearance of pRBC would be affected by cell recovery *in-vivo*, with recovery of some cell properties, e.g., as induced by rejuvenation procedure, potentially variable, with other storage-induced changes being potentially irreversible [65].

It is not yet known what flow, cell-cell, and/or cell-wall interactions in circulation may be responsible for pRBC degradation post transfusion, nor their respective potential extents, nor which *in-vitro* MF testing regimes/conditions may best simulate the relevant physiological stresses that induce hemolysis. Bead milling with cylindrical beads had been shown to generate several different types of mechanical stress in liquid medium, thus allowing testing of RBC MF under substantially different conditions [59, 64]. In this study, MF of each sample was tested using several different approaches. The difference in relative success of each approach may be indicative of its suitability for probing for RBC storage lesion. Additionally, identification of the differences in flow stresses generated through each of the methods of stress application may offer insight on the nature of pRBC membrane structural elements most affected by storage and *in-vivo* cell recovery.

Proper accounting for the multiple-unit transfusions is one of the limitations of this study. While the initial protocol stipulated single-unit transfusions, inclusion of double-unit events as well turned out to be operationally unavoidable. However, when more than one unit is transfused, the results are averaged both in terms of unit functional properties and in per-unit transfusion outcomes. In general, referring to a transfusion procedure as transfusion of a “unit” of pRBC may be somewhat misleading, because in addition to variability of unit Hb concentration, the volume of individual units can significantly vary – even if they have been manufactured according to the same method. Adjustments made for average blood volume for the three pRBC unit types used in the study improved predictive capability of all correlations involving transfusion-associated change in Hb (data not shown). Such adjustment, however, would not fully address the unit volume variability issue, and tracking of actual transfused volume would be desirable for any study attempting to use changes in patient Hb as a possible outcome metric. Moreover, the need for an additional unit to be transfused could be a response to patient condition (e.g., more severe anemia or blood loss) and/or potentially a response to an inadequate patient response to the first unit, as determined by the physician. Both possibilities could introduce a bias into the study that has not been accounted for.

Blood collection in the study protocol was aimed at obtaining a post-transfusion blood sample as close as possible to 24 hours after the transfusion. While effort was made to follow this timing, actual adherence turned out to be low – resulting in substantial variation (1 hour to 7 days, although most were within 1 day) in post-transfusion blood collection times, and the exact time of post-transfusion blood draw being impossible to determine. It is known that the majority (up to 75%) of the hemolysis of transfused pRBC occurs within 24 hours of transfusion. Thus, variation in the time of post-transfusion blood draw could have a significant effect on both primary and secondary outcomes as measured by this study, and variability in the time of draw is another limitation on the presented study.

Some consented patients, who received blood transfusion at sufficiently significant intervals, participated more than once (about 35% of the subjects). This introduced repeat sampling into the data analysis. This approach could provide valuable data on intra-patient variably of transfusion outcomes, but was not optimal here due to a limited sample size. Variable intervals between repeat transfusions likely added additional variability to outcomes. In all cases, the between-transfusions interval was smaller than typical RBC survival time, which would add a variable in itself of distribution of transfused RBC ages as of the time of collection from the donor [39] with the addition of variable unit time in storage at transfusion.

Overall, this study supports the need for further investigating functional properties of pRBC as predictors of cell hemodynamic function when in circulation and thus of transfusion success of improving oxygen delivery. RBC Mechanical Fragility (MF), as represented by certain MF profile-based parameters employed here, was shown to be a significant predictor of transfusion-associated changes in patient hemoglobin concentration. Despite the results being negatively affected by several factors that were not fully accounted for in the study, RBC MF by itself predicted up to 15% of changes in transfusion-associated hemoglobin variability. Inclusion of the number of units transfused showed the potential to improve predictive capability (up to 30% predictive value of the primary outcome), potentially reflecting the importance of accounting for patient condition. It also was shown that the means of applying the bead-induced mechanical stress affects the predictive value of the MF results. It could be expected that optimization of stress application parameters to better target the clinical application of assessing storage-lesion-associated cell damage with MF would improve predictive capability of the metrics.

## Data Availability

All data produced in the present study are available upon reasonable request to the authors

## Acknowledgements

This work was funded by a Phase 1 SBIR grant from NIH/NHLBI (#1R43HL121865-01A1) as a collaborative project by Blaze Medical Devices and the University of Michigan. The authors gratefully acknowledge the contributions of Randall Bath of Blaze Medical Devices for his work on assay performance, the employees of the University of Michigan Blood Bank and specifically Blood Bank administrative manager Theresa Downs for their assistance with the study, and Patrick Hines of Functional Fluidics for helpful discussions during reanalysis of the data, following the initial work.

